# Trends, patterns, determinants and socio-economic inequality of high-risk fertility behaviour (HRFB) among Bangladeshi women: evidence from Bangladesh Demographic Health Survey

**DOI:** 10.64898/2026.08.04.26359654

**Authors:** Raisha Binte Islam, Syed Toukir Ahmed Noor

## Abstract

**Background:** Globally and in Bangladesh, high-risk fertility behaviour (HRFB) continues to be a significant public health issue, contributing to negative health outcomes for both mothers and children. So, this study aimed to evaluate the trends, prevalence, determinants, and socio-economic inequalities associated with HRFB among presently married women of reproductive age by utilising data from the Bangladesh Demographic and Health Survey (BDHS).

**Methods:** We analysed data from 19,060 currently married women aged 15–49 years. HRFB was defined as the presence of any of the following: maternal age (<18 or >34 years), short birth intervals (<24 months), or high birth order (≥4). Two outcome variables were constructed: a binary indicator for any HRFB (yes/no) and a three-category variable indicating no, single, or multiple HRFBs. Bivariate analysis was conducted to determine the prevalence of HRFB, and multilevel mixed-effect logistic and multinomial regression models were applied to identify determinants, accounting for the complex survey design. Socioeconomic inequalities were examined using concentration indices and concentration curves.

**Results:** Overall, 58% of women experienced at least one HRFB, with 33% exhibiting a single HRFB and 25% multiple HRFBs. Women who married after age 18 years, had higher education, were exposed to media, or had educated husbands were significantly less likely to experience HRFB. Higher odds of HRFB were associated with rural residence, lower household wealth, and regions such as Mymensingh, Barishal, and Chattogram. Significant inequalities were observed, with HRFB disproportionately concentrated among women with lower wealth (CIX = −0.427, p<0.001) and no education (CIX = −0.268, p<0.001).

**Conclusion:** In Bangladesh, HRFB remains prevalent and is unevenly distributed across socio-economic and geographic groups. Targeted interventions aimed at delaying early marriage, improving educational attainment for women and their partners, expanding mass media outreach, and increasing access to reproductive healthcare-especially among disadvantaged and rural populations-are essential for reducing HRFB and improving maternal health outcomes.

## Introduction

Over the past two decades, a notable global reduction in maternal mortality has occurred. Between 2000 and 2020, the worldwide maternal mortality ratio (MMR) decreased by 34%, from 339 to 223 deaths per 100,000 live births [1] and the global under-5 mortality rate has dropped by 59%, from 93 deaths per 1000 live births in 1990 to 37 per 1000 live births in 2023 [2]. Despite these advancements, maternal and child mortality are still major public health issues, especially in low-and middle-income nations. An analysis of data from the Demographic and Health Survey (DHS) across 45 countries revealed a noteworthy positive correlation between deaths and malnutrition among children under the age of five and specific fertility-related behavioural risk factors [3]. High-risk fertility behaviour (HRFB), one major contributor to these adverse outcomes, is a set of reproductive patterns that increase the likelihood of complications for both mothers and their children. These behaviours typically include childbearing at a very young (<18) or advanced age (>35), short birth intervals, and high-parity births [4,5]. Maternal HRFB is considered a significant bio-demographic risk factor that hinders efforts to reduce maternal and child morbidity and mortality [6,7]. Given these associations, addressing and reducing high-risk fertility behaviour has become a public health priority, crucial for improving maternal and child nutrition and for further decreasing mortality rates.

It is essential to emphasise that HRFBs, whether they occur independently or in combination, are the primary underlying causes of adverse health outcomes for both the mother and her infant. The empirical evidence has established a correlation between childbearing at either a younger or older age and an increased risk of stillbirths, preterm births, and neonatal fatalities [8–12]. Young maternal age is also associated with higher rates of child malnutrition, including stunting, wasting, and low birth weight [9,10,13], while older maternal age is linked to a greater risk of genetic anomalies, pregnancy complications, and cesarean deliveries [14]. Additionally, research has shown a higher risk of maternal mortality at both younger and older ages; indeed, a “J”-shaped correlation between maternal mortality and age has been found [15]. When considering birth intervals, short birth intervals are associated with increased risks of adverse infant health outcomes like preterm birth, low birth weight, and early neonatal death [16]. Moreover, children born from short-interval pregnancies may face developmental and health issues, and the mother may face maternal anaemia [17–20]. Additionally, high-parity births are associated with a greater likelihood of maternal death and undernutrition among children [21,22]. These findings underscore the severe consequences of HRFB for both maternal and child health worldwide.

Globally, HRFB continues to be a significant public health concern, especially in settings with pronounced socioeconomic disparities. Limited access to education, healthcare, and reproductive services contributes significantly to HRFB, particularly in low-and middle-income countries. Studies have shown that women with higher education levels tend to delay childbirth, have fewer children, and space births appropriately, while those with little or no education are more likely to experience early childbirth, high parity, and short interbirth intervals [23,24]. The wealth index is a powerful predictor of HRFB since it includes several economic status markers. Research shows that compared to women from poorer homes, those from wealthier households often have fewer children, longer birth intervals, and better access to healthcare and family planning services [25,26]. Several studies have been undertaken to ascertain the levels and determinants of high-risk fertility behaviour in several developing nations, including the Democratic Republic of the Congo (DRC), Ethiopia, and Bangladesh [25,27,28]. In Bangladesh, the maternal mortality ratio is 156 per 100,000 live births, and neonatal mortality stands at 20 per 1,000 live births [29]. A study found that 67.7% of women experienced HRFB, with 22.1% exposed to multiple risks [28]. Factors such as age, religion, type of delivery and pregnancy, and contraceptive use were significantly associated with HRFB [27,28].

Although there are several studies worldwide to support the evidence of different exposures to high-risk fertility behaviours as a high-priority maternal and child health concern, very few studies in Bangladesh have evaluated HRFB in women of reproductive age comprehensively. Therefore, to inform effective prevention strategies and policy interventions, a comprehensive investigation into the determinants and broader landscape of high-risk fertility behaviour among Bangladeshi women is urgently needed. To the best of our knowledge, this is the first study in Bangladesh to use the most recent nationally representative data from the Bangladesh Demographic and Health Survey (BDHS) to simultaneously examine the trends, patterns, determinants, and socio-economic inequalities of HRFB within a single analysis. While previous studies have largely focused on the consequences of HRFB for maternal and child health, few have explored the full spectrum of underlying factors or changes over time [30]. By addressing this critical evidence gap, our study offers valuable insights to guide targeted, equity-driven interventions aimed at reducing HRFB and improving reproductive health outcomes in Bangladesh.

## Methods

### Data source and sampling

The study primarily used data from the 2022 Bangladesh Demographic and Health Survey (BDHS), conducted between August and December 2022, while earlier BDHS rounds (1997–98 to 2017–18) were additionally used to describe trends in the prevalence of HRFB. In BDHS 2022, a two-stage stratified sample approach was utilized in BDHS 2022, with the Bangladesh Bureau of Statistics (BBS) constructing the sampling frame drawn from the 2011 Population Census. Using probability proportional to size, 675 enumeration areas (EAs), comprising 237 urban and 438 rural, were first selected. The second step, when 45 households per EA were chosen to provide reliable demographic and health data for both urban and rural populations, was based on the household listings inside these EAs. The final report of the BDHS 2022 offers more information on the sampling approach [31].

### Population and Sample Size

The BDHS 2022 sampled 30,330 households nationwide, of which 30,149 were occupied and approached. A total of 30,018 households were successfully interviewed, resulting in a household response rate of 99.6%. In the interviewed households, 30,358 ever-married women aged 15–49 were identified as eligible for individual interview, of whom 20,217 were eligible for the full questionnaire, and 10,141 were eligible for the short questionnaire; interviews were completed with 30,078 women overall, yielding a response rate of 99.1%. Widowed women (n = 866), divorced (n = 413), or separated/not living with their husband (n = 262) were excluded, as our analysis focuses on currently married women. A further 9,550 women were excluded due to missing information on key variables; the majority of this exclusion reflects the survey’s sub-sampling design, whereby husband/partner characteristics (husband’s age, education, and occupation) and age at marriage are collected only from women administered the full questionnaire, so women who received the short questionnaire are structurally missing these items. After these exclusions, 19,060 (weighted) currently married women aged 15–49 years were retained for the analysis.

### Study variables and measurement

#### Outcome variables

The primary outcome variable in this study was maternal HRFB. Following the BDHS definition, HRFB was determined based on the respondent’s risk status if she were to conceive at the time of the survey [31]. Women were classified as having HRFB if they met any of the following criteria: being younger than 18 years or older than 34 years, having a birth interval of less than 24 months, or having a birth order of four or higher.

For analysis, HRFB was operationalised in two distinct ways. First, HRFB status was defined as a binary outcome indicating whether the woman had any HRFB (coded as 1 for at least one high-risk characteristic and 0 for none). Second, HRFB level was defined as a three-category outcome capturing the intensity of risk: no HRFB (no high-risk behaviour), single HRFB (presence of one high-risk behaviour), and multiple HRFB (presence of two or more high-risk behaviours). This dual specification allowed us to examine both the overall occurrence of HRFB and gradients in risk severity [7,25,32].

#### Explanatory variables

We developed a brief conceptual framework (Fig 1) based on existing literature on high-risk fertility behaviour, which suggests that maternal sociodemographic characteristics, household context, and community environment may influence HRFB. Guided by this framework, explanatory variables were selected from prior studies and grouped into these domains to examine their association with HRFB [7,32]. The selected sociodemographic and economic variables included in the analysis are: age at first marriage (<18 and ≥18 years), respondent’s education (no education, primary, secondary, and higher), religion (non-Muslim and Muslim), mass media exposure (no and yes), current use of contraceptive methods (yes and no), husband’s age (15–30, 31–34, 35–39, 40–44, 45–49, and 50+ years), husband’s education (no education, primary, secondary, and higher), number of household members (≤4 and 5 or more), wealth index (poorest, second, middle, fourth, and richest), place of residence (urban and rural), and administrative division (Barishal, Chattogram, Dhaka, Khulna, Mymensingh, Rajshahi, Rangpur, and Sylhet).

**Fig 1:**
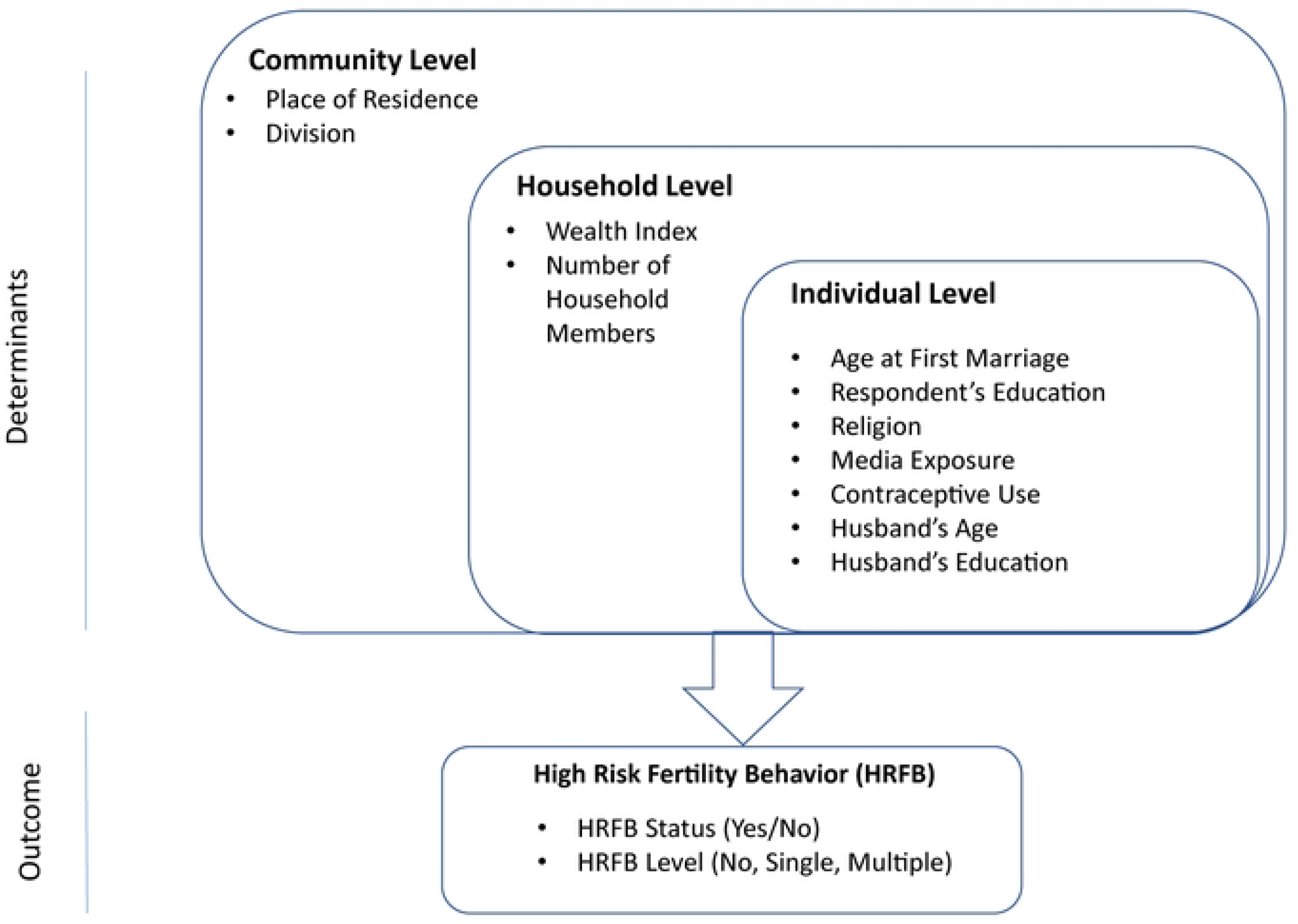
Conceptual Framework of HRFB.

#### Statistical analysis

Data cleaning, recoding, and analysis were conducted using Stata version 17.0 (StataCorp LP, College Station, Texas). Following the DHS guidelines, Sample weights were applied, and adjustments were made to accommodate the complex survey design, accounting for primary sampling units (PSUs) and strata to ensure national representativeness. The Stata “Svyset” command was utilised to handle the complex survey design [33]. The analysis adhered to the STROBE cross-sectional reporting guidelines [34]. ArcGIS version 10.8 was used to display the division-wise distribution of HRFB, and a divisional shapefile was employed to prepare the map layout. The shapefile was obtained from the Humanitarian Data Exchange (HDX), a platform managed by the United Nations Office for the Coordination of Humanitarian Affairs (available at https://data.humdata.org/dataset/cod-ab-bgd).

Exploratory data analysis was conducted to examine the characteristics of the dataset, using frequencies and percentages for categorical variables, along with 95% confidence intervals (CIs). Chi-squared tests were applied to categorical variables in the bivariate analysis. Given the large sample size and the categorical nature of the variables, this test was suitable for examining associations; expected cell frequencies were reviewed to ensure that the main assumptions were not violated. Due to the hierarchical structure of the data, we used multilevel mixed-effects logistic regression models for the binary outcome variable, any HRFB, and the findings were presented as 95% CIs and adjusted odds ratios (AORs). Models included random intercepts at the cluster (PSU) level to capture unobserved heterogeneity between clusters, while covariate effects were treated as fixed. For HRFB level (no, single, and multiple), a multinomial logistic regression model was used to assess associations, and the results were presented as relative risk ratios (RRRs) with corresponding 95% CIs. To evaluate model performance and the contribution of the multilevel specification, we reported intraclass correlation coefficients (ICCs) and model fit statistics, including the Akaike Information Criterion (AIC) and the Bayesian Information Criterion (BIC), for the multilevel logistic models. Multicollinearity was evaluated using the Variance Inflation Factor (VIF), and the variable ‘husband’s age’ was excluded from the model due to evidence of high collinearity (VIF = 6.72). In the final model, covariates exhibited acceptable collinearity, with a mean VIF of 1.83 (maximum VIF = 3.26; minimum VIF = 1.03) [37]. To assess the impact of this exclusion, we conducted a sensitivity analysis re-estimating the model with husband’s age included (**Supplementary Table S1**). Estimates for all covariates remained materially unchanged in magnitude and direction. A p-value less than 0.05 was considered statistically significant. Complete case analysis was performed, and participants with missing data on any of the study variables were excluded from the analytical model.

Socioeconomic inequalities in HRFB were assessed using concentration curves and concentration indices (CIX) [35]. A concentration curve graphically represents the distribution of HRFB across socioeconomic strata by plotting the cumulative proportion of the population, ranked by factors like wealth or education, against the cumulative proportion of HRFB cases [36]. The 45-degree diagonal, or equality line, signifies an absence of inequality. In wealth-based analyses, curves below this line indicate that HRFB disproportionately affects wealthier women, reflecting systemic advantages in reproductive health access for affluent groups. Conversely, curves above the line suggest higher HRFB prevalence among poorer women, highlighting disparities disadvantaging lower-income populations. Similarly, for education-based inequality, curves below the equality line imply HRFB is concentrated among women with higher education, while curves above the line signal higher HRFB rates among those with no formal education.

The index takes a value between −1 and 1, with 0 indicating perfect equity. The CIX was calculated using the “convenient covariance” formula proposed by O’Donnell et al. [36], as follows:

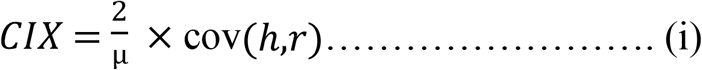

Where h represents the variable of the health sector, μ = mean of these variables, and r denotes the fractional rank of individual i (where, 1 ≤ i ≤ N; 1 = poorest/No education, N = richest/higher). Using the “lorenz” [37] and “conindex” [38] Stata commands, we computed the Concentration curve and CIX, respectively.

### Ethical Consideration

This study used de-identified secondary data from the DHS websites (https://www.dhsprogram.com/data/available-datasets.cfm/). The BDHS protocol received ethical clearance from the ICF Institutional Review Board and the Bangladesh Medical Research Council (BMRC). In the original survey, trained interviewers obtained informed consent from all respondents after explaining the purpose, procedures, duration, potential risks and benefits, and the voluntary nature of participation; respondents could decline any question or test or withdraw at any time. For participants younger than 18 years, consent was obtained from a parent or legal guardian in addition to the adolescent’s assent. The datasets available to researchers are fully anonymised and contain no personally identifiable information; therefore, no additional ethical approval was required for this secondary analysis, and permission to use the data was obtained from the DHS Program.

## Results

### Background characteristics

**Table 1** presents the demographic and socioeconomic characteristics of Bangladeshi married women included in the study. Educational attainment varied, with 47.4% having secondary education and 13.2% reporting no formal education. The majority of participants were Muslim (90.4%), and over half (58.3%) had some form of media exposure. A substantial proportion of women (67.5%) were married before the age of 18. The age distribution of husbands ranged from 15 to over 50 years, with 20.9% aged 50 or older. Regarding their education, 32.5% of husbands had completed secondary schooling. A majority of the respondents (71.7%) resided in rural areas. Regionally, the highest proportion of participants came from the Dhaka division (25.3%), followed by Chattogram (18.7%) and Rajshahi (13.2%).

**Table 1:** Background characteristics of currently married women in Bangladesh. Note: Percentages are weighted using survey sample weights to account for the complex survey design. The full analytic sample is n=19,060; minor variation in summed n across reflects the rounding of weighted frequencies, and does not indicate differences in the underlying analytic sample.

| Variable | Weighted frequency | Weighted percentage |
| --- | --- | --- |
| **Respondent's education** |  |  |
| No education | 2474 | 12.98 |
| Primary | 4907 | 25.75 |
| Secondary | 9038 | 47.42 |
| Higher | 2640 | 13.85 |
| **Religion** |  |  |
| Non-Muslim | 1833 | 9.62 |
| Muslim | 17222 | 90.35 |
| Missing | 5 | 0.03 |
| **Media Exposure** |  |  |
| No | 7940 | 41.66 |
| Yes | 11120 | 58.34 |
| **Age at first marriage** |  |  |
| <18 | 12859 | 67.47 |
| >=18 | 6201 | 32.53 |
| **Currently using contraceptive method** |  |  |
| No | 6866 | 36.02 |
| Yes | 12193 | 63.98 |
| **Husband's age** |  |  |
| 15-29 | 3484 | 18.28 |
| 30-34 | 2830 | 14.85 |
| 35-39 | 3356 | 17.61 |
| 40-44 | 3004 | 15.76 |
| 45-49 | 2398 | 12.58 |
| 50+ | 3986 | 20.92 |
| **Husband's education** |  |  |
| No education | 4121 | 21.62 |
| Primary | 5386 | 28.26 |
| Secondary | 6196 | 32.51 |
| Higher | 3356 | 17.61 |
| **Number of household members** |  |  |
| <=4 | 9065 | 47.56 |
| 5 or more | 9995 | 52.44 |
| **Wealth index** |  |  |
| Richest | 3930 | 20.62 |
| Fourth | 3991 | 20.94 |
| Middle | 3930 | 20.62 |
| Second | 3846 | 20.18 |
| Poorest | 3363 | 17.64 |
| **Place of Residence** |  |  |
| Urban | 5385 | 28.25 |
| Rural | 13675 | 71.75 |
| **Divison** |  |  |
| Barishal | 1153 | 6.05 |
| Chattogram | 3559 | 18.67 |
| Dhaka | 4817 | 25.27 |
| Khulna | 2281 | 11.97 |
| Mymensingh | 1450 | 7.61 |
| Rajshahi | 2521 | 13.23 |
| Rangpur | 2197 | 11.53 |
| Sylhet | 1082 | 5.68 |

### Trends and prevalence of HRFB

**Fig 2** demonstrates the overall trends in HRFB (single and multiple) prevalence for about three decades from 1993-94 to 2022. The prevalence has shown a mixed pattern over the years, but in the final year, the prevalence of high-risk births has declined from 66% in 1993-94 to 58% in 2022. Single HRFB has been reduced by only 1% (34 to 33%), whereas multiple HRFB has been reduced by 6% (32 to 25%).

**Fig 2:**
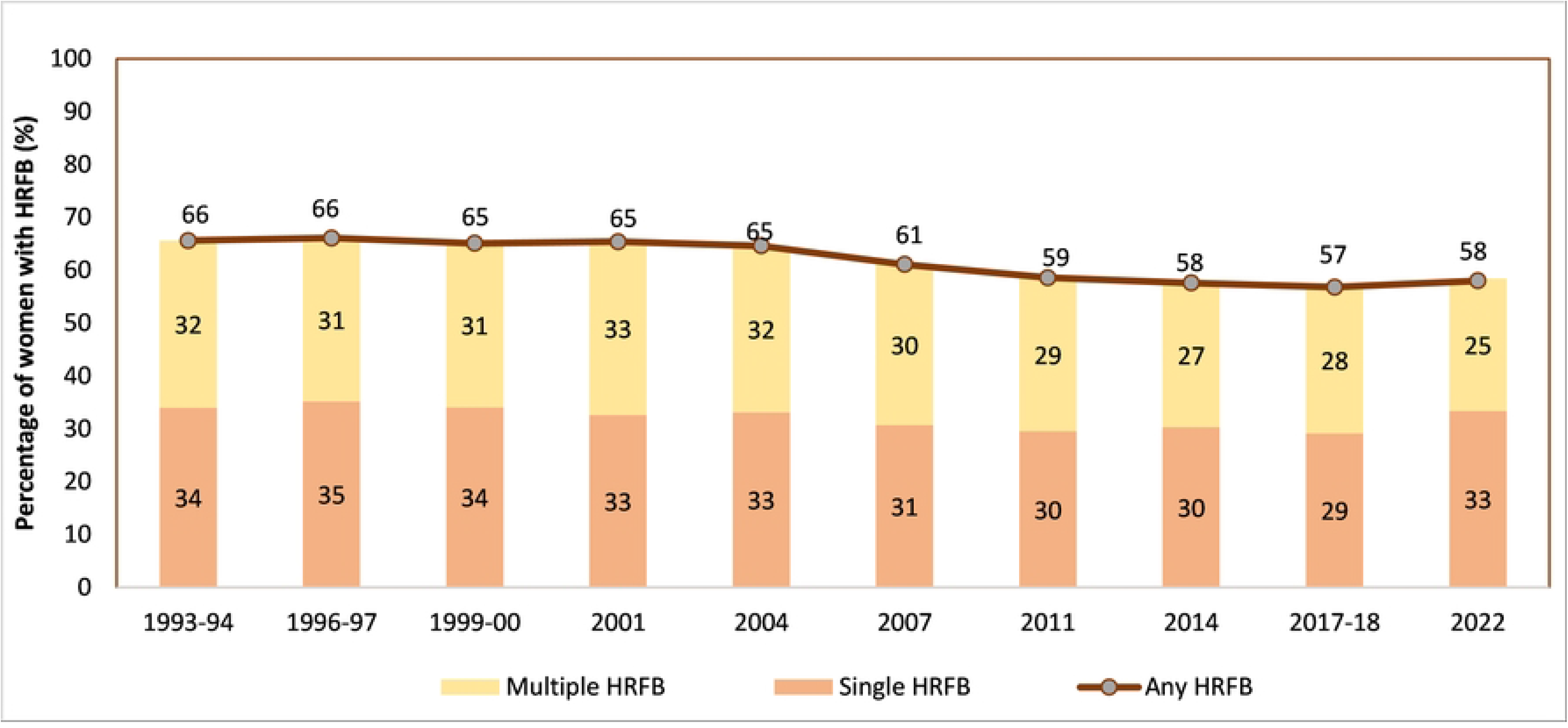
Trends in the prevalence of high-risk fertility behaviours among currently married women in Bangladesh.

**Fig 3:**
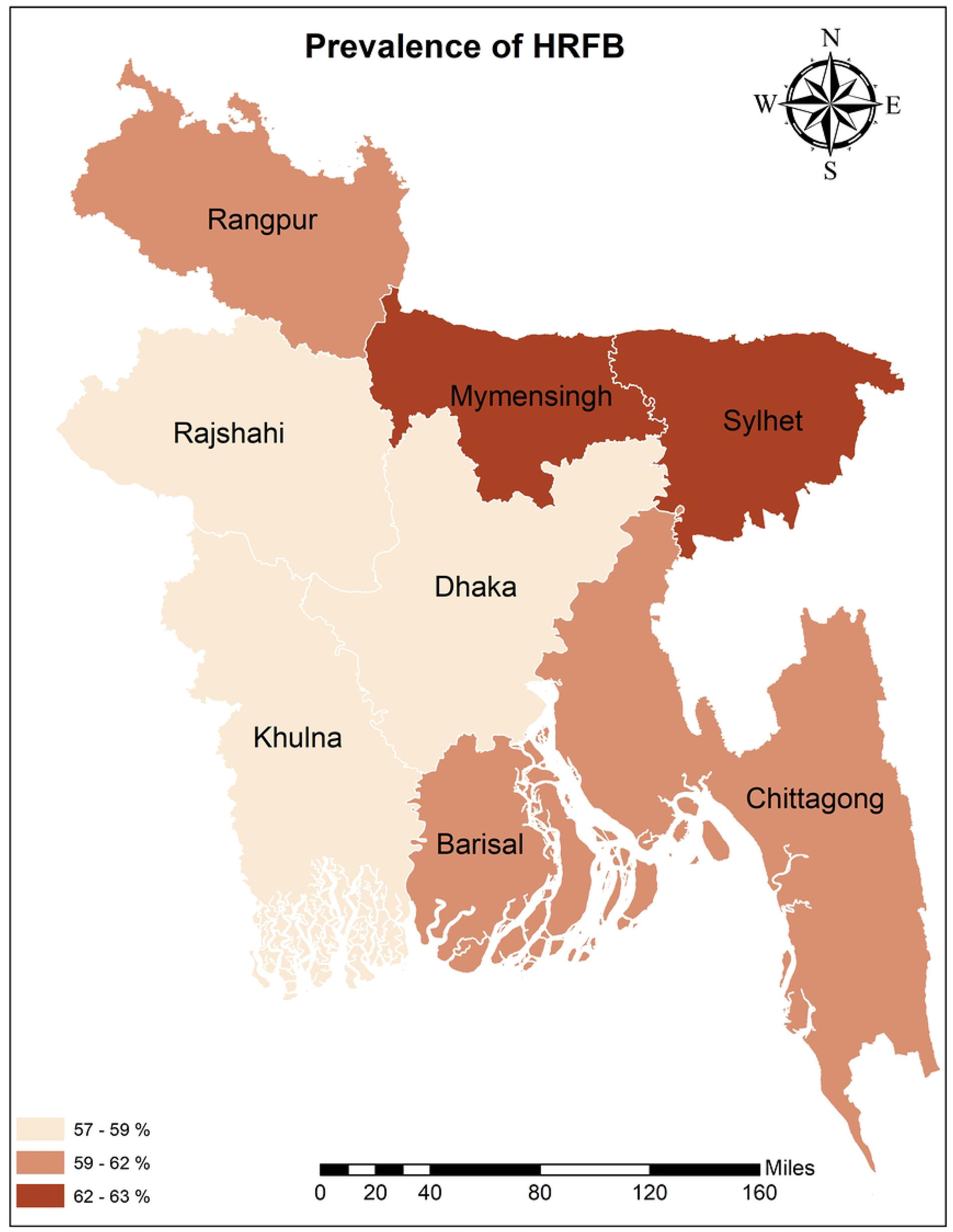
Prevalence of HRFB by administrative division of Bangladesh.

Scenario from the most recent data revealed that the most common single high-risk category was mother’s age greater than 34 years (16.4%), followed by birth interval less than 24 months (8.25%) and birth order greater than 3 (6.76%). In the case of multiple HRFB, the most frequent combination is age at birth >34 & birth order >3, followed by birth interval <24 months & birth order >3 (**Table 2**).

**Table 2:**
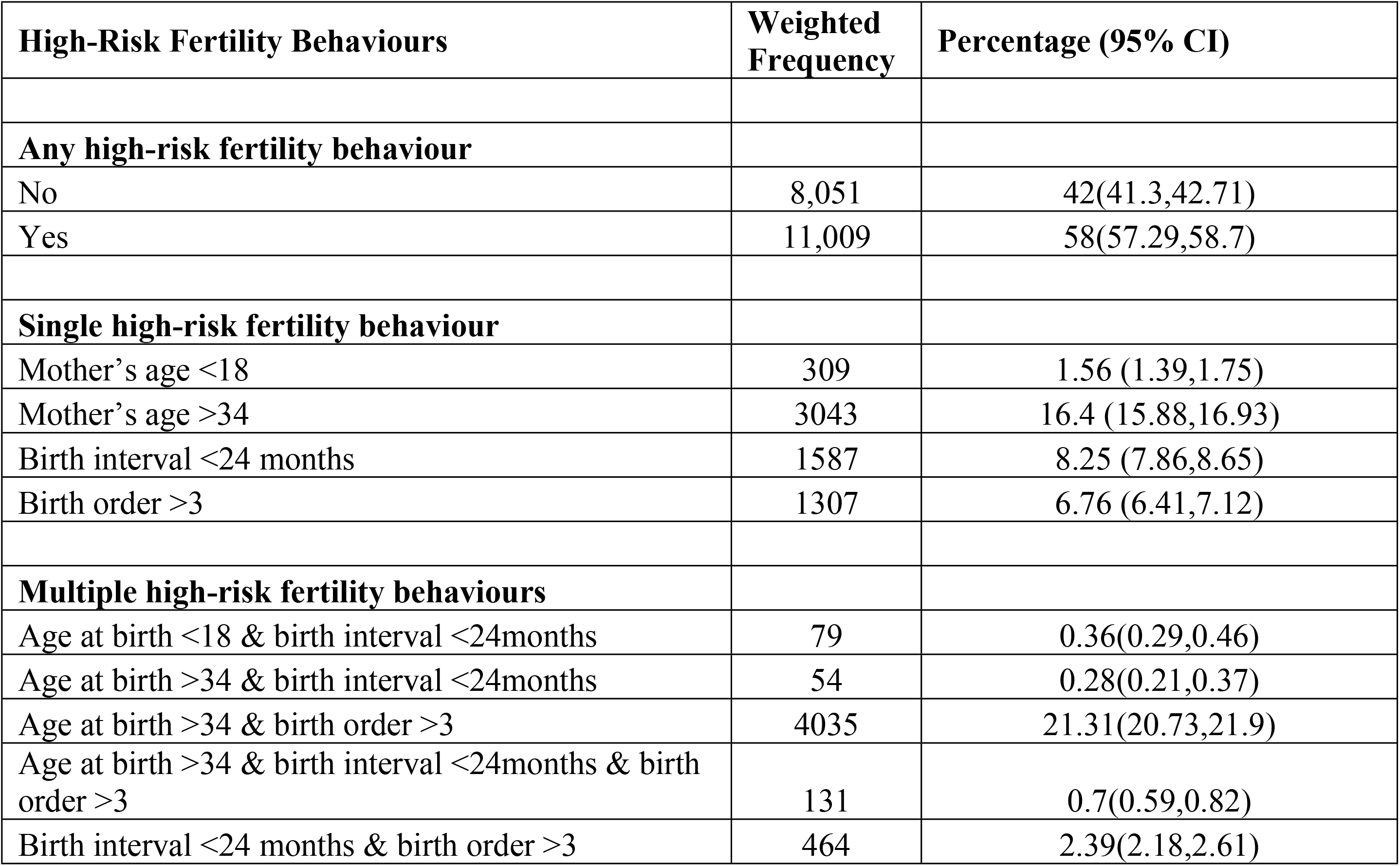
Levels of high-risk fertility behaviours among currently married women in Bangladesh.

| High-Risk Fertility Behaviours | Weighted Frequency | Percentage (95% CI) |
| --- | --- | --- |
| **Any high-risk fertility behaviour** |  |  |
| No | 8,051 | 42(41.3,42.71) |
| Yes | 11,009 | 58(57.29,58.7) |
| **Single high-risk fertility behaviour** |  |  |
| Mother's age <18 | 309 | 1.56 (1.39,1.75) |
| Mother's age >34 | 3043 | 16.4 (15.88,16.93) |
| Birth interval <24 months | 1587 | 8.25 (7.86,8.65) |
| Birth order >3 | 1307 | 6.76 (6.41,7.12) |
| **Multiple high-risk fertility behaviours** |  |  |
| Age at birth <18 & birth interval <24months | 79 | 0.36(0.29,0.46) |
| Age at birth >34 & birth interval <24months | 54 | 0.28(0.21,0.37) |
| Age at birth >34 & birth order >3 | 4035 | 21.31(20.73,21.9) |
| Age at birth >34 & birth interval <24months & birth order >3 | 131 | 0.7(0.59,0.82) |
| Birth interval <24 months & birth order >3 | 464 | 2.39(2.18,2.61) |

### Prevalence of HRFB by background characteristics

**Table 3** represents the distribution of HRFB across selected demographic and socio-economic characteristics. The results of chi-square tests revealed that almost all the selected background variables were significantly associated with HRFB. Respondents who married after age 18 exhibited the highest prevalence (35.9%) of single HRFB. Multiple HRFB was higher (55.7%) in the women’s group, where they had no education, followed by primary (37.3%) and secondary (17.5%), whereas single HRFB was higher in the higher education group (36.7%). Muslim women had higher exposure to multiple HRFB (27.1%), whereas non-muslim women had higher exposure to single HRFB (40.9%). Women who were not exposed to the media had a higher rate of multiple HRFB (32.7%). Women who were using a contraceptive method had a slightly greater prevalence of HRFB (58.5%). Consistent with women’s educational attainment, HRFB prevalence was higher among women whose husbands had no formal education. Women living in the larger family exhibited greater HRFB prevalence (62.6%). Therefore, women coming from the lowest wealth quantile had the highest prevalence of multiple HRFB (32.2%). ‘

**Table 3:**
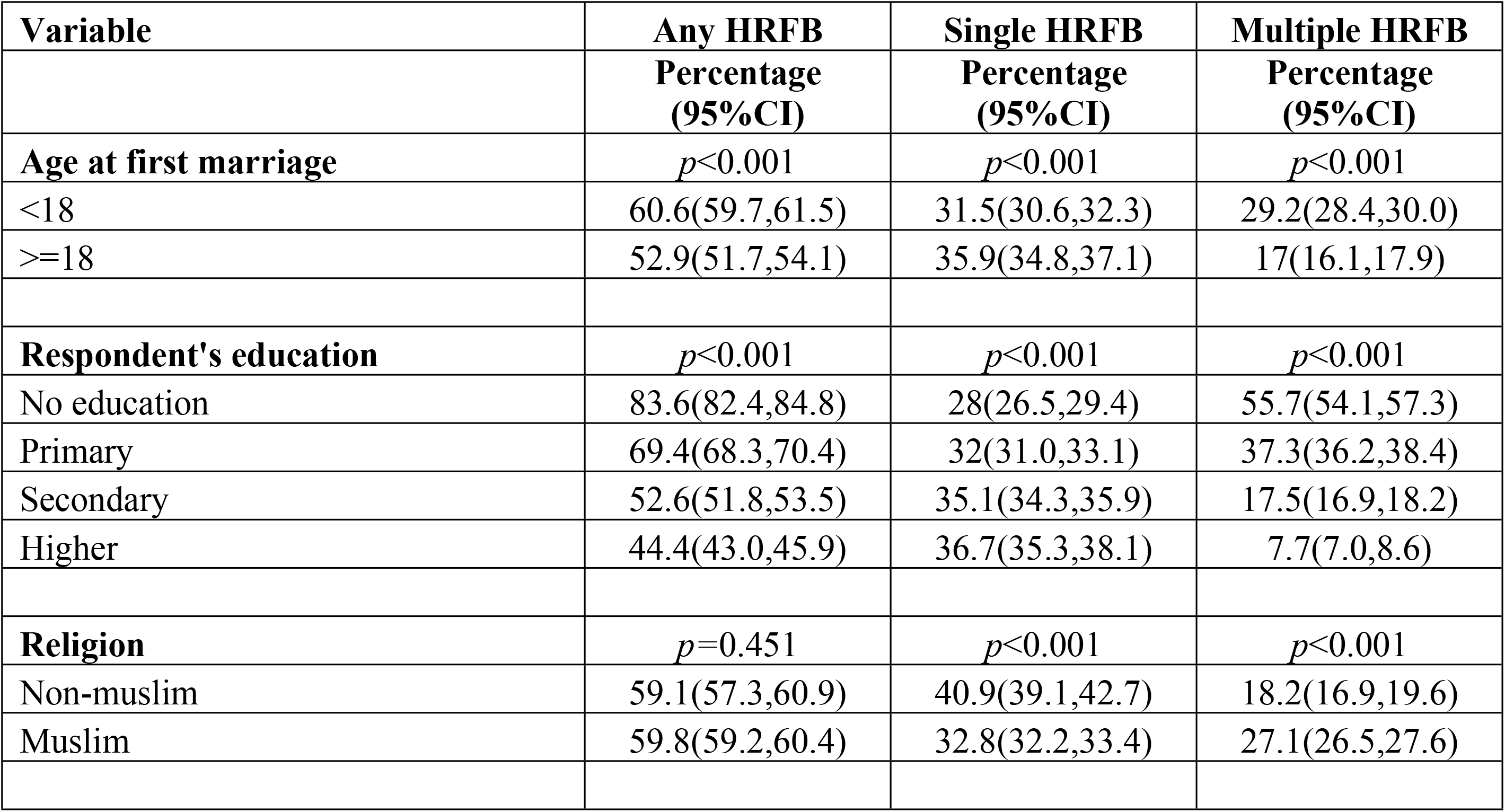
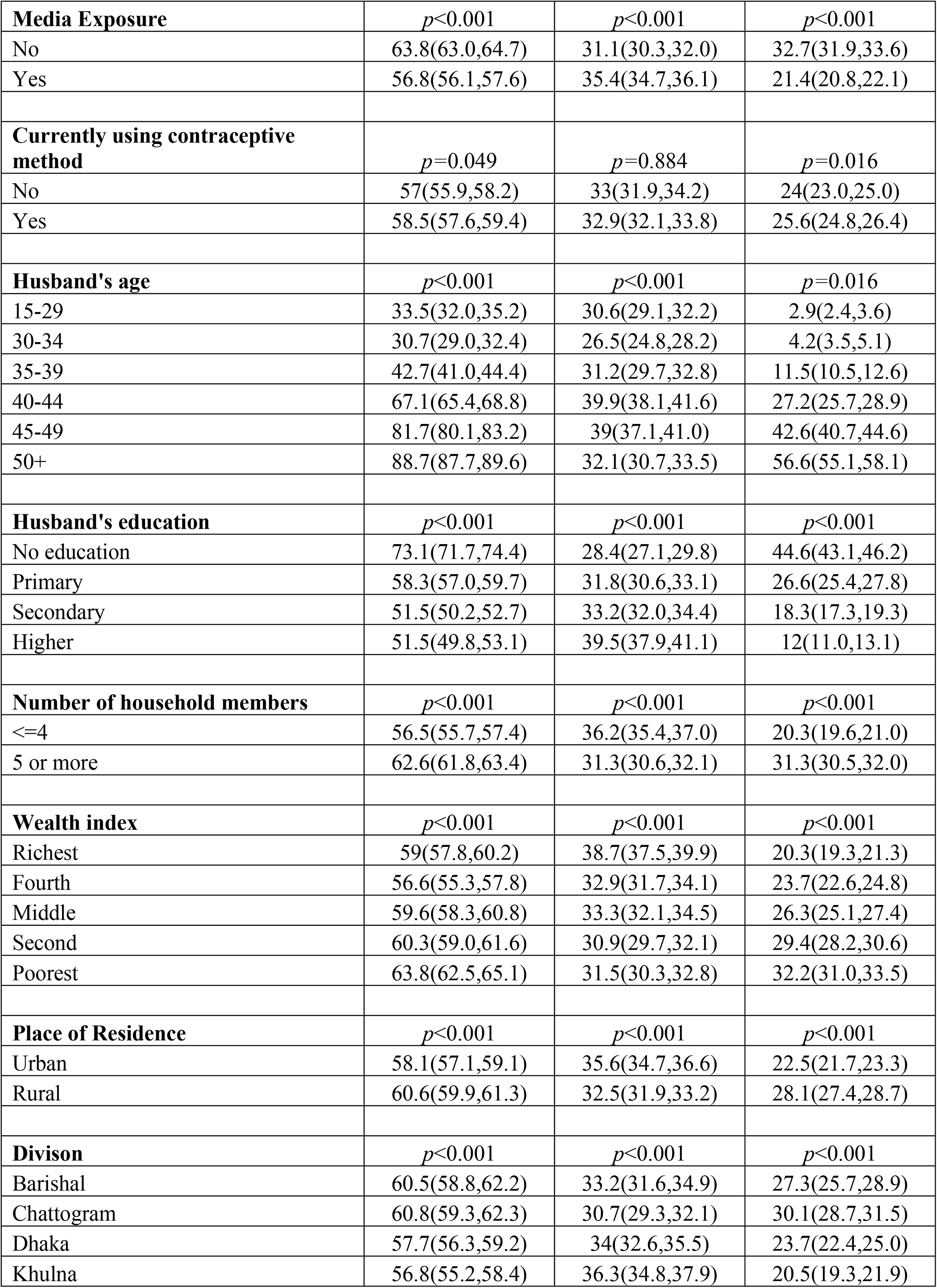
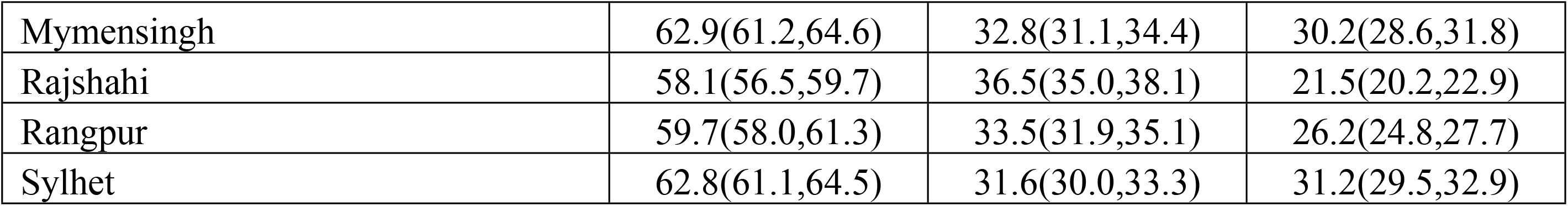
Distribution of high-risk fertility behaviours by background characteristics among currently married women in Bangladesh. Regarding place of residence, HRFB was more prevalent among women residing in rural areas and in the divisions of Mymensingh, Sylhet, Chattogram, and Barishal (**Fig 3**).

| Variable | Any HRFB<br>Percentage<br>(95%CI) | Single HRFB<br>Percentage<br>(95%CI) | Multiple HRFB<br>Percentage<br>(95%CI) |
| --- | --- | --- | --- |
| **Age at first marriage** | $p<0.001$ | $p<0.001$ | $p<0.001$ |
| <18 | 60.6(59.7,61.5) | 31.5(30.6,32.3) | 29.2(28.4,30.0) |
| >=18 | 52.9(51.7,54.1) | 35.9(34.8,37.1) | 17(16.1,17.9) |
| **Respondent's education** | $p<0.001$ | $p<0.001$ | $p<0.001$ |
| No education | 83.6(82.4,84.8) | 28(26.5,29.4) | 55.7(54.1,57.3) |
| Primary | 69.4(68.3,70.4) | 32(31.0,33.1) | 37.3(36.2,38.4) |
| Secondary | 52.6(51.8,53.5) | 35.1(34.3,35.9) | 17.5(16.9,18.2) |
| Higher | 44.4(43.0,45.9) | 36.7(35.3,38.1) | 7.7(7.0,8.6) |
| **Religion** | $p=0.451$ | $p<0.001$ | $p<0.001$ |
| Non-muslim | 59.1(57.3,60.9) | 40.9(39.1,42.7) | 18.2(16.9,19.6) |
| Muslim | 59.8(59.2,60.4) | 32.8(32.2,33.4) | 27.1(26.5,27.6) |
| **Media Exposure** | $p<0.001$ | $p<0.001$ | $p<0.001$ |
| No | 63.8(63.0,64.7) | 31.1(30.3,32.0) | 32.7(31.9,33.6) |
| Yes | 56.8(56.1,57.6) | 35.4(34.7,36.1) | 21.4(20.8,22.1) |
| **Currently using contraceptive method** | $p=0.049$ | $p=0.884$ | $p=0.016$ |
| No | 57(55.9,58.2) | 33(31.9,34.2) | 24(23.0,25.0) |
| Yes | 58.5(57.6,59.4) | 32.9(32.1,33.8) | 25.6(24.8,26.4) |
| **Husband's age** | $p<0.001$ | $p<0.001$ | $p=0.016$ |
| 15-29 | 33.5(32.0,35.2) | 30.6(29.1,32.2) | 2.9(2.4,3.6) |
| 30-34 | 30.7(29.0,32.4) | 26.5(24.8,28.2) | 4.2(3.5,5.1) |
| 35-39 | 42.7(41.0,44.4) | 31.2(29.7,32.8) | 11.5(10.5,12.6) |
| 40-44 | 67.1(65.4,68.8) | 39.9(38.1,41.6) | 27.2(25.7,28.9) |
| 45-49 | 81.7(80.1,83.2) | 39(37.1,41.0) | 42.6(40.7,44.6) |
| 50+ | 88.7(87.7,89.6) | 32.1(30.7,33.5) | 56.6(55.1,58.1) |
| **Husband's education** | $p<0.001$ | $p<0.001$ | $p<0.001$ |
| No education | 73.1(71.7,74.4) | 28.4(27.1,29.8) | 44.6(43.1,46.2) |
| Primary | 58.3(57.0,59.7) | 31.8(30.6,33.1) | 26.6(25.4,27.8) |
| Secondary | 51.5(50.2,52.7) | 33.2(32.0,34.4) | 18.3(17.3,19.3) |
| Higher | 51.5(49.8,53.1) | 39.5(37.9,41.1) | 12(11.0,13.1) |
| **Number of household members** | $p<0.001$ | $p<0.001$ | $p<0.001$ |
| $\leq 4$ | 56.5(55.7,57.4) | 36.2(35.4,37.0) | 20.3(19.6,21.0) |
| 5 or more | 62.6(61.8,63.4) | 31.3(30.6,32.1) | 31.3(30.5,32.0) |
| **Wealth index** | $p<0.001$ | $p<0.001$ | $p<0.001$ |
| Richest | 59(57.8,60.2) | 38.7(37.5,39.9) | 20.3(19.3,21.3) |
| Fourth | 56.6(55.3,57.8) | 32.9(31.7,34.1) | 23.7(22.6,24.8) |
| Middle | 59.6(58.3,60.8) | 33.3(32.1,34.5) | 26.3(25.1,27.4) |
| Second | 60.3(59.0,61.6) | 30.9(29.7,32.1) | 29.4(28.2,30.6) |
| Poorest | 63.8(62.5,65.1) | 31.5(30.3,32.8) | 32.2(31.0,33.5) |
| **Place of Residence** | $p<0.001$ | $p<0.001$ | $p<0.001$ |
| Urban | 58.1(57.1,59.1) | 35.6(34.7,36.6) | 22.5(21.7,23.3) |
| Rural | 60.6(59.9,61.3) | 32.5(31.9,33.2) | 28.1(27.4,28.7) |
| **Divison** | $p<0.001$ | $p<0.001$ | $p<0.001$ |
| Barishal | 60.5(58.8,62.2) | 33.2(31.6,34.9) | 27.3(25.7,28.9) |
| Chattogram | 60.8(59.3,62.3) | 30.7(29.3,32.1) | 30.1(28.7,31.5) |
| Dhaka | 57.7(56.3,59.2) | 34(32.6,35.5) | 23.7(22.4,25.0) |
| Khulna | 56.8(55.2,58.4) | 36.3(34.8,37.9) | 20.5(19.3,21.9) |
| Mymensingh | 62.9(61.2,64.6) | 32.8(31.1,34.4) | 30.2(28.6,31.8) |
| Rajshahi | 58.1(56.5,59.7) | 36.5(35.0,38.1) | 21.5(20.2,22.9) |
| Rangpur | 59.7(58.0,61.3) | 33.5(31.9,35.1) | 26.2(24.8,27.7) |
| Sylhet | 62.8(61.1,64.5) | 31.6(30.0,33.3) | 31.2(29.5,32.9) |

### Determinants of HRFB

**Table 4** represents the results of the multinomial analysis. Model 1 is a binomial logistic regression model that presents the adjusted odds of any HRFB. In contrast, Model 2 is a multinomial logistic regression model that provides relative risk ratios for single and multiple HRFB among women across all selected socio-economic and demographic variables. In both models, we considered no HRFB as the reference category.

**Table 4:** Determinants of high-risk fertility behaviour among currently married women in Bangladesh. *p < 0.10, **p < 0.05, ***p < 0.001; Ref= Reference Category; AOR= Adjusted Odds Ratio; RRR=Relative Risk Ratio; HRFB= High Risk Fertility Behaviour.

|  | **Model 1 (Binomial Logistic)** | **Model 2 (Multinomial Logistic)** |  |
| --- | --- | --- | --- |
|  | **Any HRFB vs No HRFB** | **Single HRFB vs No HRFB** | **Multiple HRFB vs No HRFB** |
| **Variable** | **AOR (95% CI)** | **RRR (95% CI)** | **RRR (95% CI)** |
| **Age at first marriage** |  |  |  |
| <18 | Ref | Ref | Ref |
| >=18 | 0.91 (0.85, 0.98)* | 1.04(0.96, 1.13) | 0.71(0.65, 0.79) *** |
| **Respondent's education** |  |  |  |
| No education | Ref | Ref | Ref |
| Primary | 0.54 (0.48, 0.61)*** | 0.72(0.63, 0.83) *** | 0.44(0.38, 0.50) *** |
| Secondary | 0.29 (0.25, 0.32)*** | 0.52(0.45, 0.60) *** | 0.15(0.13, 0.17) *** |
| Higher | 0.18 (0.15, 0.20)*** | 0.36(0.30, 0.43) *** | 0.06(0.04, 0.07) *** |
| **Religion** |  |  |  |
| Non-muslim | Ref | Ref | Ref |
| Muslim | 0.96 (0.87, 1.07) | 0.81(0.73, 0.90) *** | 1.45(1.25, 1.67) *** |
| **Media Exposure** |  |  |  |
| No | Ref | Ref | Ref |
| Yes | 0.86 (0.80, 0.92)*** | 0.94(0.87, 1.01) | 0.75(0.69, 0.82) *** |
| **Currently using contraceptive method** |  |  |  |
| No | Ref | Ref | Ref |
| Yes | 1.05 (0.98, 1.11) | 1.02(0.95, 1.09) | 1.1(1.01, 1.19) * |
| **Husband's education** |  |  |  |
| No education | Ref | Ref | Ref |
| Primary | 0.69 (0.63, 0.76)*** | 0.83(0.74, 0.92) *** | 0.58(0.52, 0.64) *** |
| Secondary | 0.62 (0.56, 0.69)*** | 0.78(0.70, 0.88) *** | 0.48(0.42, 0.54) *** |
| Higher | 0.82 (0.72, 0.93)** | 1.05(0.91, 1.20) | 0.59(0.49, 0.69) *** |
| **Number of HH** |  |  |  |
| <=4 | Ref | Ref | Ref |
| 5 or more | 1.27 (1.19, 1.35)*** | 1.01(0.95, 1.09) | 1.86(1.72, 2.02) *** |
| **Wealth index** |  |  |  |
| Richest | Ref | Ref | Ref |
| Fourth | 0.91 (0.84, 0.97)** | 0.80(0.74,0.87)*** | 1.10(1.00,1.21)* |
| Middle | 1.02 (0.95,1.10) | 0.87(0.80,0.95)** | 1.31(1.19,1.44)*** |
| Second | 1.06 (0.98, 1.14) | 0.83(0.76,0.90)*** | 1.50(1.36,1.64)*** |
| Poorest | 1.22 (1.13, 1.132)*** | 0.92(0.85,1.00) | 1.80(1.64,1.97)*** |
| **Place of Residence** |  |  |  |
| Urban | Ref | Ref | Ref |
| Rural | 1.08 (1.01, 1.16)* | 1.04(0.96, 1.12) | 1.19(1.08, 1.30) *** |

| Divison |  |  |  |
| --- | --- | --- | --- |
| Dhaka | Ref | Ref | Ref |
| Barishal | 1.28 (1.13, 1.44)*** | 1.15(1.00, 1.32) * | 1.53(1.31, 1.79) *** |
| Chattogram | 1.15 (1.03, 1.28)* | 1(0.88, 1.13) | 1.44(1.24, 1.66) *** |
| Khulna | 1.02 (0.91, 1.15) | 1.09(0.97, 1.24) | 0.89(0.76, 1.04) |
| Mymensingh | 1.34 (1.18, 1.52)*** | 1.3(1.13, 1.49) *** | 1.41(1.20, 1.66) *** |
| Rajshahi | 1.06 (0.94, 1.18) | 1.15(1.01, 1.30) * | 0.91(0.77, 1.06) |
| Rangpur | 1.12 (1.00, 1.27) | 1.11(0.97, 1.26) | 1.16(0.99, 1.36) |
| Sylhet | 1.04 (0.92, 1.18) | 0.97(0.84, 1.12) | 1.16(0.98, 1.35) |
| ICC (SE) | 0.14 (0.04) |  |  |
| Log-likelihood | -12174.8 | -18810.5 |  |
| AIC | 24400 | 37717 |  |
| BIC | 24596 | 38094 |  |

\* $p < 0.10$ , \*\* $p < 0.05$ , \*\*\* $p < 0.001$ ; Ref= Reference Category; AOR= Adjusted Odds Ratio;

Model 1 reveals several significant associations between sociodemographic factors and HRFB among women. Women who married after the age of 18 were 9% less likely to experience HRFB compared to those who married before 18 years of age (AOR=0.91; 95% CI=0.85, 0.98). Education emerged as a strong protective factor; women with higher levels of education had substantially lower odds of HRFB than those with no formal education (AOR=0.18; 95% CI=0.15, 0.20). Women exposed to media were 14% less likely to experience HRFB than those without media exposure (AOR=0.86; 95% CI=0.80, 0.92). Similarly, the educational attainment of husbands influenced HRFB outcomes; women whose husbands had higher education were 18% less likely to have HRFB than those whose husbands had no education (AOR=0.82; 95% CI=0.72, 0.93). The wealth gradient was strongly evident, with the poorest households having significantly higher odds of HRFB than the richest households (AOR=1.22; 95% CI=1.13, 1.32). Rural residents were associated with 8% higher odds than their urban counterparts (AOR=1.08; 95% CI=1.01, 1.16). Finally, regional disparities were evident, with women from Mymensingh, Barishal, and Chattogram divisions exhibiting higher odds of HRFB than those from Dhaka, highlighting geographic variations in fertility behaviour.

Similar to model 1, model 2 demonstrates the associations between various background characteristics and the relative risk of experiencing single and multiple HRFB compared to the no-risk group. The relative risk of multiple HRFB was significantly reduced among women who married after the age of 18, with a relative risk ratio (RRR) of 0.71 (RRR=0.71; 95% CI=0.65, 0.79). Education appeared as a key factor, with the relative risks for both single and multiple HRFB significantly decreased in the higher education cohort compared to the illiterate cohort. Religious affiliation also played a role, as Muslim women exhibited a 45% higher relative risk of multiple HRFB than their non-Muslim counterparts (RRR = 1.45; 95% CI: 1.25, 1.67). In contrast, the relative risk of experiencing multiple HRFB is 25% lower in women who were exposed to media (RRR=0.75; 95% CI=0.69, 0.82). Women whose husbands had attained higher education had a significantly lower risk of multiple HRFB (RRR = 0.59; 95% CI = 0.49, 0.69), highlighting the importance of educational attainment in both partners. Household size was another contributing factor; respondents from larger families had a higher relative risk of both single and multiple HRFB compared to those from smaller households. Moreover, women from the poorest wealth quintile had an 80% higher relative risk of experiencing multiple HRFB than those from the richest group (RRR=1.80; 95% CI=1.64, 1.97), and it was statistically significant. Rural residence was consistently linked to a higher relative risk of both single and multiple HRFB when compared to urban residence, and lastly, women from Barishal, Chattogram, and Mymensingh had 53%, 44%, and 41% higher relative risk, respectively, than women from Dhaka.

### Socio-economic inequality of HRFB

**Fig 4 (A & B)** shows the difference in HRFB across different wealth groups and different educational qualification groups among reproductive-aged women in Bangladesh. The concentration curves showed a significant wealth-and education-related inequality since they consistently appeared above the equality line. Furthermore, the CIX values provided additional evidence for this disparity, with CIX = −0.427 (p-value <0.001) for wealth and CIX = −0.268 (p-value <0.001) for education. These negative CIX values imply that women from the lower wealth index and the illiterate group were more likely to have HRFB.

**Fig 4:**
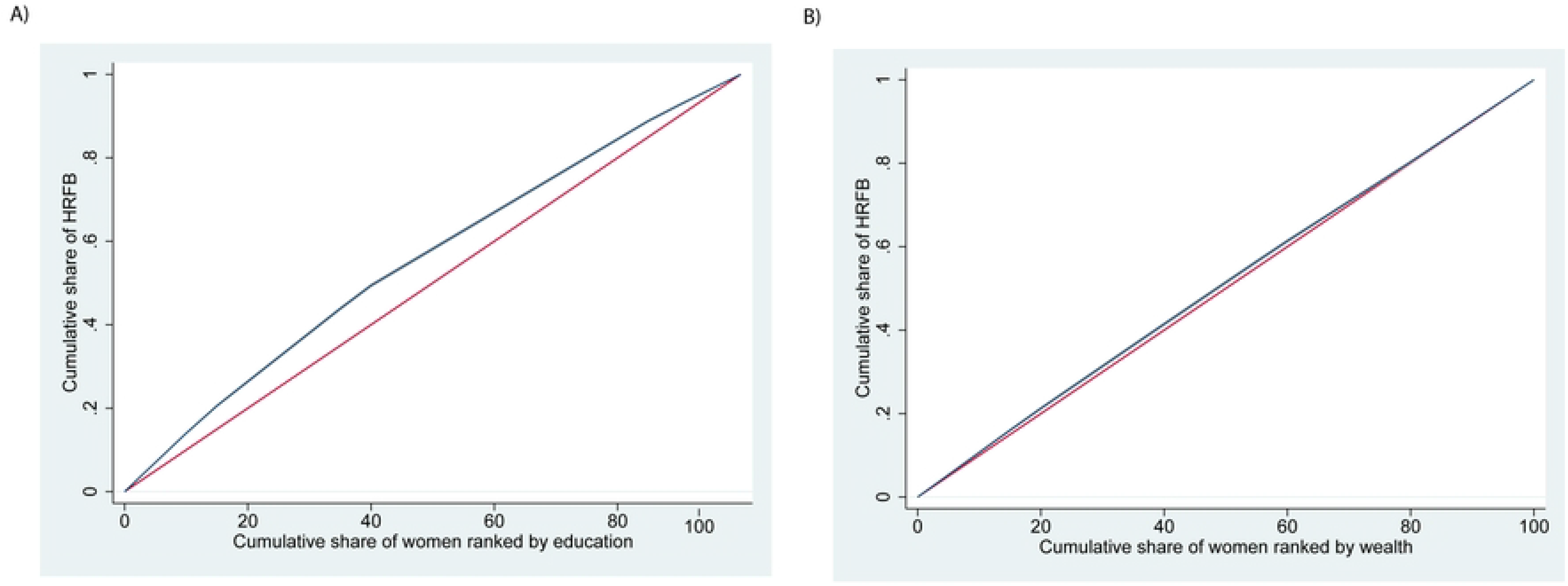
Socio-economic inequality among currently married women in Bangladesh.

## Discussion

The main goal of this study was to examine the trends, patterns, determinants, and socio-economic inequalities of HRFB among reproductive-aged women in Bangladesh. Though HRFB has declined slightly over the past three decades, still, every six out of ten married women in Bangladesh are experiencing high-risk births. According to the regression analysis, higher education, media exposure, better wealth status, and urban residence reduce the risk of HRFB. However, inequality was evident; HRFB was found to be disproportionately concentrated among poorer and less educated women.

HRFB trend in Bangladesh reveals that the prevalence of HRFB remained relatively stable and high from 1993-94 through 2004, with about two-thirds of women experiencing HRFB. A notable decline began after 2004, with the percentage dropping steadily to its lowest point in 2017-18 [39]. However, this downward trend reversed slightly in 2022, with the prevalence rising again according to our analysis. This increase should be interpreted with caution but may reflect a combination of factors, including post-pandemic disruptions in healthcare access and family planning services, as well as broader socioeconomic challenges during the pandemic [31]. These types of disruptions have been reported in other studies examining the effects of COVID-19 on reproductive health and related services [40–42]. This variation underscores the need for resilient health systems and sustained efforts to maintain progress in reducing HRFB, particularly in the context of public health crises [43].

Education was found to be a key determinant for HRFB in our study; women with higher levels of education and an educated husband had a substantially lower likelihood of experiencing a high-risk birth. A similar finding was evident in previous studies in sub-Saharan African countries, including Ethiopia and Kenya, with higher education levels associated with reduced HRFB [32,44,45]. Educated partners usually have higher reproductive health literacy, enabling informed decisions about birth timing, spacing, and family size. Educated couples are more likely to recognise the risks of short interpregnancy intervals (<24 months) and prioritise optimal maternal age for childbirth. Moreover, education encourages a desire for smaller families, perhaps because of conflicting priorities like career commitments and the aspiration to allocate resources toward fewer children. A trend reflected in Bangladesh, where women with ≥10 years of schooling have 1.8 children on average, compared to 3.2 among those without formal education [31].

Our study also found that women who married after the age of 18 had a significantly lower chance of experiencing HRFB. Early marriage is a well-documented risk factor for HRFB because it often leads to early childbearing, shorter birth intervals, and higher parity, all of which increase maternal and child health risks [46]. Marrying after 18 increases women’s chances for education and economic involvement and helps them to reach more physical maturity, hence empowering their knowledge and autonomy in family planning and fertility choices. These results align with previous studies demonstrating that adolescent marriage is associated with increased risks of maternal morbidity and mortality, as well as poor neonatal outcomes [6,47]. Furthermore, delayed marriage is frequently linked to higher contraceptive use and greater reproductive decision-making power, factors that contribute to lowering the likelihood of HRFB [46]. Therefore, interventions aimed at preventing child marriage and promoting girls’ education and empowerment remain critical strategies to reduce HRFB and improve maternal and child health outcomes.

Our findings reveal significant geographic disparities in HRFB, with rural residents exhibiting higher odds of HRFB compared to their urban counterparts. This finding is consistent with existing studies conducted in Ethiopia and Congo, which highlight persistent rural-urban disparities in HRFB [47,48]. Rural areas often face challenges such as limited access to quality healthcare services, lower levels of education, and socioeconomic disadvantages, all of which contribute to higher fertility risks. In Bangladesh, rural women typically have reduced access to family planning resources and lower contraceptive use, which increases the likelihood of early childbearing, short birth intervals, and higher parity [28,49,50]. Cultural norms and gender dynamics common in rural areas may further limit women’s control over reproductive choices, hence increasing these dangers.

Regional disparities were also evident in our analysis, with women from the Mymensingh, Barishal, and Chattogram divisions exhibiting higher odds of HRFB compared to those from Dhaka. These geographic variations reflect the uneven distribution of healthcare infrastructure, socioeconomic development, and educational attainment across Bangladesh. Similar patterns have been observed in other low-and middle-income countries, where certain regions demonstrate higher fertility risks due to limited health service coverage and entrenched socio-cultural practices [45,48]. Furthermore, regions like Barishal and Chattogram have been reported to have lower contraceptive prevalence and higher rates of early marriage, which contribute to increased HRFB prevalence [43]. The differences highlight the importance of region-specific initiatives tackling the particular cultural and structural obstacles women in different regions experience.

Our study found that Muslim women had a significantly higher likelihood of experiencing multiple HRFB compared to their non-Muslim counterparts, consistent with prior research in Bangladesh and other low-and middle-income countries. The finding was also consistent with previous studies conducted in Bangladesh, which reported that Muslim women were more likely to have HRFB compared to non-Muslims [28]. This association should not be interpreted as religion itself being causative, but rather as religious affiliation potentially serving as a proxy for broader contextual factors, including community norms, gender dynamics, and structural barriers to care. Prior work suggests that in some communities, misinterpretation of religious guidance on family planning, together with social expectations around early marriage, high desired family size, and son preference, may influence contraceptive use and fertility behaviour [7,51]. Moreover, our models may not fully capture unmeasured confounders such as local service availability, quality of counselling, and neighborhood-level norms, so the observed association should be interpreted with caution. Similar patterns have been observed in other contexts, such as Kenya and Ethiopia, where Muslim women also demonstrate higher odds of HRFB [44,52].

Wealth-related inequality was evident in our study, with women from poorer households exhibiting significantly higher odds of HRFB than those from wealthier households. This finding is consistent with evidence from Bangladesh and other low-and middle-income countries, where socioeconomic disadvantage strongly correlates with increased HRFB prevalence [28,44,48]. Previous studies in Bangladesh found that HRFB was disproportionately concentrated among the poorest women, who often face barriers such as limited access to quality maternal health services, lower educational attainment, and reduced autonomy in reproductive decision-making [28,53]. Similarly, a study from Ethiopia demonstrates a widening pro-poor inequality in HRFB over time, with educational status, wealth index, and contraceptive use identified as key contributors to this disparity [54]. Poorer women tend to have lower contraceptive uptake and higher rates of early marriage and childbearing, which increase the risk of short birth intervals and high parity, core components of HRFB [30,54]. The compounded effects of poverty and limited healthcare access increase adverse maternal and child health outcomes associated with HRFB, including increased perinatal mortality and childhood undernutrition [30,53].

These findings have direct implications for current health and development priorities in Bangladesh. The concentration of HRFB among poorer women suggests the need for stronger equity-oriented implementation of the national family planning and maternal health agenda, particularly for women facing financial, geographic, and informational barriers to care. Targeted outreach through community health workers, improved access to affordable contraception, and integration of reproductive health counselling into primary care could help reduce these disparities. In addition, policies that support girls’ education, women’s empowerment, and timely utilisation of maternal services are essential to address the structural drivers of HRFB and to advance Bangladesh’s commitments to reducing maternal and child health inequalities.

## Strengths and limitations

This study has several notable strengths. To the best of our knowledge, it is the first comprehensive investigation in Bangladesh to simultaneously examine the trends and patterns of HRFB, its socioeconomic inequalities, and associated determinants using a nationally representative sample. The use of the most recent BDHS 2022 dataset, which employed rigorous sampling techniques, enhances the generalizability of the findings to women of reproductive age (15–49 years) across the country. Additionally, the large sample size and application of appropriate statistical techniques allowed for robust identification of significant predictors and assessment of inequality gradients across multiple subgroups. These methodological strengths provide policymakers and program planners with timely and reliable evidence for designing targeted maternal health interventions.

However, the study is not without limitations. First, as with most DHS-based studies, data are self-reported and thus subject to recall bias and social desirability bias, which may affect the accuracy of reported reproductive behaviours. Second, due to the cross-sectional nature of the survey, the study cannot establish causal relationships between HRFB and its determinants. Third, some relevant covariates, such as maternal comorbidities, nutritional status, dietary patterns, physical activity, and psychosocial factors, were not included in the BDHS and thus could not be examined, even though they may have significant implications for HRFB. Fourth, although we accounted for key sociodemographic and reproductive factors, residual confounding remains possible because we could not include all potentially relevant individual, household, and community-level variables. Relatedly, some covariates (such as husband’s age) showed evidence of multicollinearity; as a result, husband’s age was excluded from the final multivariable models, which may have led to loss of information despite sensitivity analyses suggesting that the main estimates were robust. Fifth, we relied on complete-case analysis for handling missing data, and if the data were not missing completely at random, this could bias the estimates. Finally, socioeconomic characteristics were measured at the time of the interview, which may not accurately reflect the woman’s status at the time of pregnancy or childbirth.

## Conclusion

This study reveals that HRFB remains a substantial public health challenge in Bangladesh, with the burden disproportionately concentrated among women from poorer households. Given our finding that socioeconomic disadvantage is a key driver of HRFB, there is a critical need to expand access to affordable and high-quality reproductive health services for the poorest segments of the population. Strengthening the reach of family planning, antenatal, and maternal health programs in economically disadvantaged communities will be vital for reducing these inequities. Our results also demonstrate that the educational attainment of women and their husbands is strongly associated with lower odds of HRFB. This underscores the importance of promoting girls’ education and preventing early marriage through targeted policies and community engagement. By investing in education, women are better equipped to make informed reproductive choices and delay childbearing, thereby reducing their risk of high-risk fertility patterns.

The study further highlights that rural residence and living in certain divisions, such as Mymensingh, Barishal, and Chattogram, are linked to higher HRFB compared to urban areas and the Dhaka division. To address these geographic disparities, it is essential to strengthen healthcare infrastructure and outreach in rural and high-risk regions, ensuring that tailored family planning and maternal health interventions are accessible to women regardless of where they live. Finally, the association between religious affiliation and HRFB, with Muslim women exhibiting higher odds of multiple HRFB, points to the need for culturally sensitive health education strategies. Engaging religious and community leaders in reproductive health promotion can help address misconceptions and encourage healthy fertility behaviours within all communities.

Overall, strengthening rural health systems, improving access to reproductive health services, and promoting community-based education programs tailored to regional contexts are essential to reducing HRFB and improving maternal and child health outcomes in Bangladesh.

## Declarations

## Data Availability

This study used secondary data from the 2022 Bangladesh Demographic and Health Survey (BDHS). The BDHS 2022 datasets are available from the DHS Program for legitimate research purposes upon registration and approval at https://www.dhsprogram.com/data/available-datasets.cfm. Because we do not own these data, we are not permitted to republish the individual-level dataset. However, the complete Stata do-file used for data cleaning, variable construction, weighting, and all statistical analyses is provided as Supporting Information (**S1 File**), enabling full reproducibility of our results for researchers who obtain access to the BDHS 2022 data from the DHS Program.

## Acknowledgements

We are grateful to the DHS team for allowing us to conduct the analysis of this study using the BDHS 2022 data set.

## Author Contributions

Conceptualization, R.B.I and S.T.A.N; Data curation, R.B.I; Formal analysis, R.B.I; Methodology, S.T.A.N; Visualization, R.B.I; Writing, R.B.I; Writing – review and editing, S.T.A.N; All authors have read and agreed to the published version of the manuscript.

## Consent for publication

Not applicable.

## Supporting information

S1 File. Stata do-file for data analysis.

S2 File. Supplementary material.

